# Risk Adjusted Non-Pharmaceutical Interventions for the Management of COVID-19 in South Africa

**DOI:** 10.1101/2020.07.15.20149559

**Authors:** Joshua Choma, Fabio Correa, Salah-Eddine Dahbi, Kentaro Hayasi, Benjamin Lieberman, Caroline Maslo, Bruce Mellado, Kgomotso Monnakgotla, Jacques Naudé, Xifeng Ruan, Finn Stevenson

## Abstract

A global analysis of the impact of non-pharmaceutical interventions (NPIs) on the dynamics of the spread of the COVID-19 indicates that these can be classified using the stringency index proposed by the Oxford COVID-19 Government Response Tracker (OxCGRT) team. The world average for the coefficient that linearises the level of transmission with respect to the OxCGRT stringency index is *α*_*s*_ = 0.01*±*0.0017 (95% C.I.). The corresponding South African coefficient is *α*_*s*_ = 0.0078 ± 0.00036 (95% C.I.), compatible with the world average. Here, we implement the stringency index for the recently announced 5-tier regulatory alert system. Predictions are made for the spread of the virus for each alert level. Assuming constant rates of recovery and mortality, it is essential to increase *α*_*s*_. For the system to remain sub-critical, the rate with which *α*_*s*_ increases should outpace that of the decrease of the stringency index. Monitoring of *α*_*s*_ becomes essential to controlling the post-lockdown phase. Data from the Gauteng province obtained in May 2020 has been used to re-calibrate the model, where *α*_*s*_ was found increase by 20% with respect to the period before lockdown. Predictions for the province are made in this light.

## 1. Introduction

Stringent lockdown measures, such as those implemented in South Africa and other countries, have been successful in curbing the spread. Epidemiological control would favor prolonging the lockdown in order to prevent the resurgence of the spread. However, stringent lockdown measures have devastating effects on the economy. As a result, the control of the epidemic needs to enter a new phase, where lockdown measures are expected to be relaxed while maintaining the rate of spread so as not to overwhelm the health care system. This would then allow for a gradual reactivation of the economy.

On the 23^*rd*^ of April 2020 President Cyril Ramaphosa announced a 5 tier COVID-19 regulatory alert system to gradually relax the stringency level of the initial response and imposed interventions. This regulatory system will be implemented in South Africa from the beginning of May 2022. ^1^ This system aims to sufficiently contain the spread whilst gradually opening all of the sectors of the economy in different alert levels. The decision of whether to open different sectors of the economy will be based on a number of complex factors including the risk of infection associated with the particular work environment and the overall economic value of the sector. The 5 tiers are enumerated *L*1 − *L*5; level one (*L*1) is the least stringent and *L*5 is the most stringent. The lock-down implemented in South Africa on the 26^*th*^ of March 2020 can be equated to *L*5.

### 1.1. On Modeling and Control

Mathematical modelling of the COVID-19 virus within South Africa is essential for making informed decisions with regards to the restrictions necessary at any given time. The decision to elevate or demote the South African Alert Level must be made with caution and with all factors considered. Predictions based on an epidemiologically informed mathematical model have been developed in a parent paper^2^. The SIRD model used therein incorporated both latent and dynamic infection characteristics of the virus. This means that the model takes into account both the asymptomatic and symptomatic people infected with the virus in the mathematics. This is crucial because there has been research that suggests that people who are asymptomatic make up a large portion of the total people infected.^3^ A detailed explanation on the country specific SIRD models and a global overview of the effectiveness of the various NPIs that have been implemented around the world can be seen in the parent paper^2^. The mathematical model’s parameters were fit to the COVID-19 time-series case data for 24 countries and 25 States of the United States and certain important parameters were extracted in order to develop predictions and make comparisons. Each parameter’s time variation was captured with a kernel function which was used to characterise the dynamic characteristics of the epidemic to changes in the host environment.

Levels of control were measured by the Oxford COVID-19 Government Response Tracker (OxCGRT) stringency index. This index, denoted *p* in our work, takes values from 0 to 100 and is indicative of the severity and number of discrete measures that have been taken to try to curb the spread of COVID-19. The measures of control of each country within the OxCGRT set are comprehensive and therefore there are more than 5 levels of control in *p*. It is possible though to characterise the 5 South African alert levels in terms of the Oxford indicators S1-S7.

## 2. Explanation of the OxCGRT Stringency Index

The OxCGRT has developed a valuable database for comparing countries response strategies. ^4 5^ The database contains the following levels of control (coded using ordinal numbers) and timing for 139 countries:

**Table 1:** Table Showing Indicators and their Corresponding Responses.

| OxCGRt Indicator | Intervention Response |
| --- | --- |
| S1: | School closure |
| S2: | Workplace closure |
| S3: | Cancel public events |
| S4: | Close public transport |
| S5: | Public information campaign |
| S6: | Domestic travel bans |
| S7: | International travel bans |

Also included in this data set is a Stringency Index^6^, *p*, in our notation, which provides a single number that captures the overall level of intervention implied by combinations of the ordinal numbers S1-S7. The Oxford stringency index is calculated using a weighted average of the above seven non-pharmaceutical interventions^6^. For each policy response measure S1-S7, OxCGRT use the ordinal value (and add one if the policy is general rather than targeted). This creates a score between 0 and 2 and for S5, and 0 and 3 for the other six responses^5^.

The OxCGRT stringency index is given by:

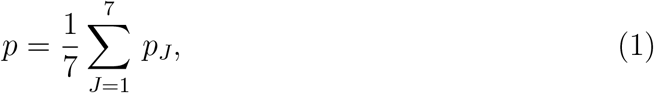

where *p*_*J*_ is defined by:

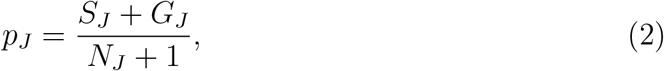

with *G*_*J*_ = 1 if the effect is general (and 0 otherwise), and *N*_*J*_ is the cardinality of the intervention measure^5,6^. In the case where there is no requirement of general vs. targeted (S7), the +1 in the denominator and the *G*_*J*_ in the numerator are omitted from the equation to form:

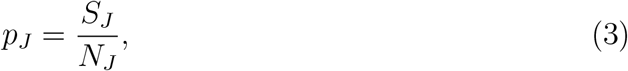

## 3. Adaption of the OxCGRT Stringency Index for the South African Alert Levels

It is necessary for the South African Alert Levels (L1-L5) to be characterised in terms of the Oxford indicators (NPIs) S1-S7 so that predictions can be made using the aforementioned model.

The calculation of a stringency index for South Africa will be similarly based on the average of all the individual intervention measures using Eq.(1). However, in South Africa it is not necessary to include the notion of general vs. targeted interventions, therefore Eq.(3) will be utilised for the calculation of individual *p*_*J*_ values; *p*_*J*_ ∈ [0, 1]. This calculation method is used in order to incorporate degrees of implementation for each NPI (S1-S7) that is appropriate for the specific South African Alert Levels.

The chosen cardinality of the interventions in a South African Context can be seen in Table 2.

**Table 2:**
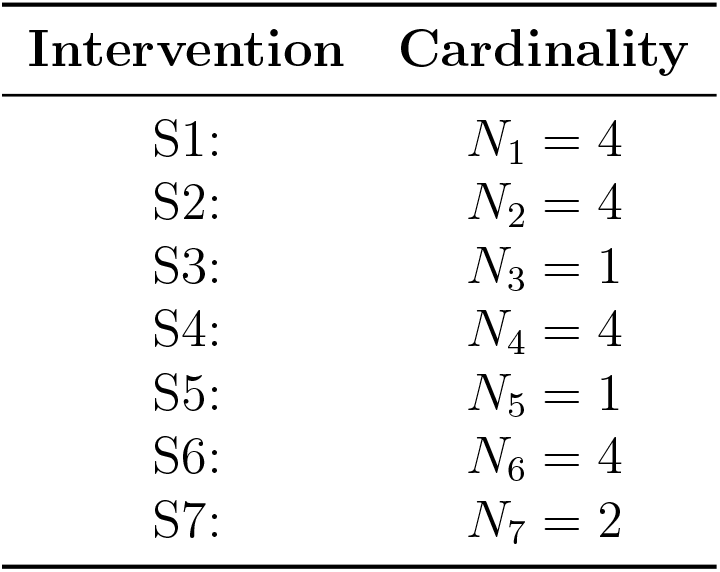
Table Showing Interventions and their Corresponding Cardinality for South Africa.

The above cardinalities were chosen based on the following criteria:

- If *N*_*J*_ = 1, there is no need for the incorporation of levels of intervention implementation in the South African context.
- If *N*_*J*_ = 2, there is a need for there to be 2 levels of intervention implementation in the South African context.
- If *N*_*J*_ = 4, there is a need for there to be 4 levels of intervention implementation in the South African context.

### 3.1. Assumptions

The following assumptions are made for S1, S3 and S5 control interventions ^7^:

- The *“School Closure” S*1 intervention will be a function of both the alert level and the stage of school reopening.
- The “Cancel Public Events” *S*3 intervention measure will be implemented for all of the South African Alert Levels. This assumption is made based on the following statement from the South African Government: *“The following restrictions will remain in place after the national lock-down, and regardless of the level of alert at any given time: Sit-in restaurants and hotels; Bars and shebeens; Conference and convention centres; Entertainment venues, including cinemas, theatres, and concerts; Sporting events; Religious, cultural and social gatherings. No gatherings of more than 10 people outside of a workplace will be permitted*.*”*
- The “Public Information Campaign” S5 intervention measure will be implemented for all of the South African Alert Levels. This assumption was made because the government should not stop distributing COVID-19 information and recommendations until the COVID-19 virus is no longer a epidemic in South Africa.

### 3.2. South African Stringency Index Results

The results reported in Table 3 were obtained using Eq. (3). This includes the cardinality values *N*_1_ - *N*_7_ and the appropriate *S*_*J*_ values that align with the level of intervention of the specific South African Alert Level. Note that the S1 indicator is not only a function of the SA Alert Level but also of the stage of school reopening in South Africa.

**Table 3:** Table Showing Indicator Contribution to the Stringency Index for different South African Alert Levels. The impact of the S1 indicator is estimated separately (see text).

|  | South African Alert Level |  |  |  |  |
| --- | --- | --- | --- | --- | --- |
|  | Level 1 | Level 2 | Level 3 | Level 4 | Level 5 |
| OxCGRT Indicator |  |  |  |  |  |
| $S1$ | — | — | — | — | — |
| $S2$ | 0 | 0.25 | 0.5 | 0.75 | 1 |
| $S3$ | 1 | 1 | 1 | 1 | 1 |
| $S4$ | 0 | 0 | 0.25 | 0.5 | 1 |
| $S5$ | 1 | 1 | 1 | 1 | 1 |
| $S6$ | 0 | 0.25 | 0.5 | 1 | 1 |
| $S7$ | 0.5 | 1 | 1 | 1 | 1 |
| Total (Excl. $S1$): | 2.5 | 3.5 | 4.25 | 5.25 | 6 |

The rationale for the scoring of the indicators S2, S4, S6 and S7 can be found in Appendix A. Explanation of the effects of the S1 indicator on the stringency index can be found in Appendix B. It should be noted that the S1 indicator is time dependent. This is driven by the staged resumption of classes announced by the Minister of Basic education. As a result, the stringency index at all levels becomes time dependent as well. This is illustrated in Figure 1, where the stringency index for each level gradually decreases as classes resume in a staged fashion.

**Figure 1:**
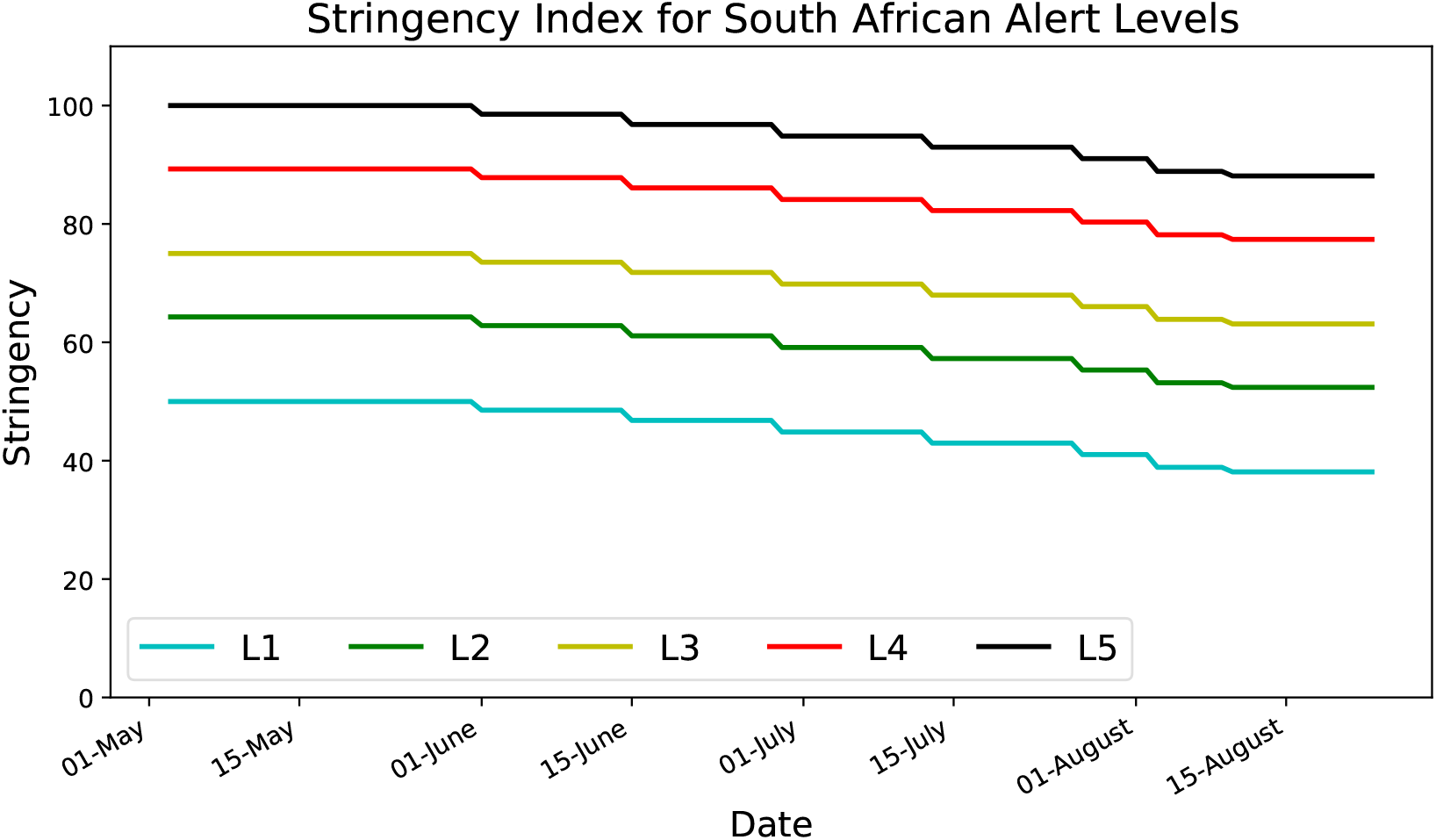
Time Varying Stringency Index for each South African Alert Levels 1-5.

### 3.3. Further Recommendations for Stringency Calculation

Further improvements could be made by more accurately quantifying the *N*_*J*_ values based on a more thorough analysis of the control criteria related to the specific interventions:

- The S2 associated cardinality *N*_2_ can be scaled based on an in-depth analysis of the businesses that are allowed to operate at each level to more accurately represent what percentage of persons are back at their workplaces the different levels.

### 3.4. Stringency Index Comparison

The achieved stringency index boundaries for the South African alert system compare favourably with those of countries around the world.

**Table 4:** Table Showing Stringency Index Boundaries for the 5 tier South African Alert System with Comparison to Countries around the World.

| Level | Achievable SA Index | Countries |
| --- | --- | --- |
| 1 | $36 < p < 50$ | Taiwan - 38 |
| 2 | $50 < p < 64$ | Sweden - 52 |
| 3 | $61 < p < 75$ | Japan - 67, Australia - 71, UK - 71 |
| 4 | $75 < p < 89$ | Canada - 86, Germany - 86 |
| 5 | $86 < p < 100$ | South Africa - 100, India - 100, Israel - 100 |

## 4. South African Predictions

The predictions performed here conform to the prescription detailed in the parent paper^2^, where two steps are followed. Firstly, the parameters of the model are extracted before and during the lockdown using South African data provided by the NICD and the Department of Health. These parameters constitute the initial conditions for the forward temporal evolution of a SIRD model. Secondly, a SIRD model is used by reasoning that the daily recovery rate and daily mortality parameter, *γ*_0_ and *d*_0_, respectively, remain constant and the transmission rate follows the functional form:

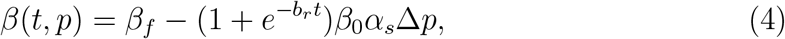

where *t* = 0 corresponds to the end of the initial lockdown period and the removal of some interventions, such that *p < p*_*max*_.

The implementation of new less restrictive measures will yield Δ*p* = *p* − *p*_*max*_ *<* 0, where *β*(*t, p*) *> β*_*f*_. Here, *β*_*f*_ is the asymptotic value of the transmission rate achieved during lockdown with *p*_*max*_, which in South Africa is considered to be equal to 100. The parameters *β*_0_, *α*_*s*_ are obtained from the data reported before and after the lockdown was enacted. Table 5 reports the parameter values used here for predictions. It should be noted that due to the specifics of the South African data the typical adjustment time, denoted 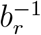, is about one day, which is to small to be attributed to a well motivated adjustment time. Several checks have been performed to ensure that this feature does not affect the stability of critical parameters, such as *α*_*s*_. Global analysis of the data indicates that 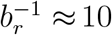 days^2^. This value is used for the temporal evolution based on Eq. 4. The value of *d*_0_ is chosen so as to achieve a mortality rate of 2.5%. The uncertainty of this estimate is 25% (68% C.I.) and it is based on a combination of inputs. This comprises a mortality matrix obtained from Wuhan data and multi-dimensional data from the Gauteng City-Region Observatory. Documenting the details of this study goes beyond the scope of this paper. The estimate used here is consistent with the current mortality rate for SA, which seems to have stabilised at 2% for the moment.

**Table 5:** Parameters obtained with South African data up until May 2^*th*^ 2020 (see text).

| $\gamma_0$ | $\beta_0$ | $\beta_f$ | $p_{max}$ | $\alpha_s$ |
| --- | --- | --- | --- | --- |
| 0.022 | 0.28 | 0.06 | 100 | 0.0078 |

Figure 2 displays the differential distributions of active cases in South Africa for different alert levels. The *x*-axis shows time in days. Results are presented in terms of the differential fractional distribution of cases. This is with respect to the total number of symptomatic individuals. If one takes 10% as the fraction of susceptible individuals that develop symptoms and test positive, one arrives at 6 million people in South Africa. This implies for Level 3 that, that about 3 million people would test positive at the apex. In practice, the number of severe cases that need hospitalisation or even ICU usage are derived from these estimates by using inputs from clinicians and practitioners.

**Figure 2:**
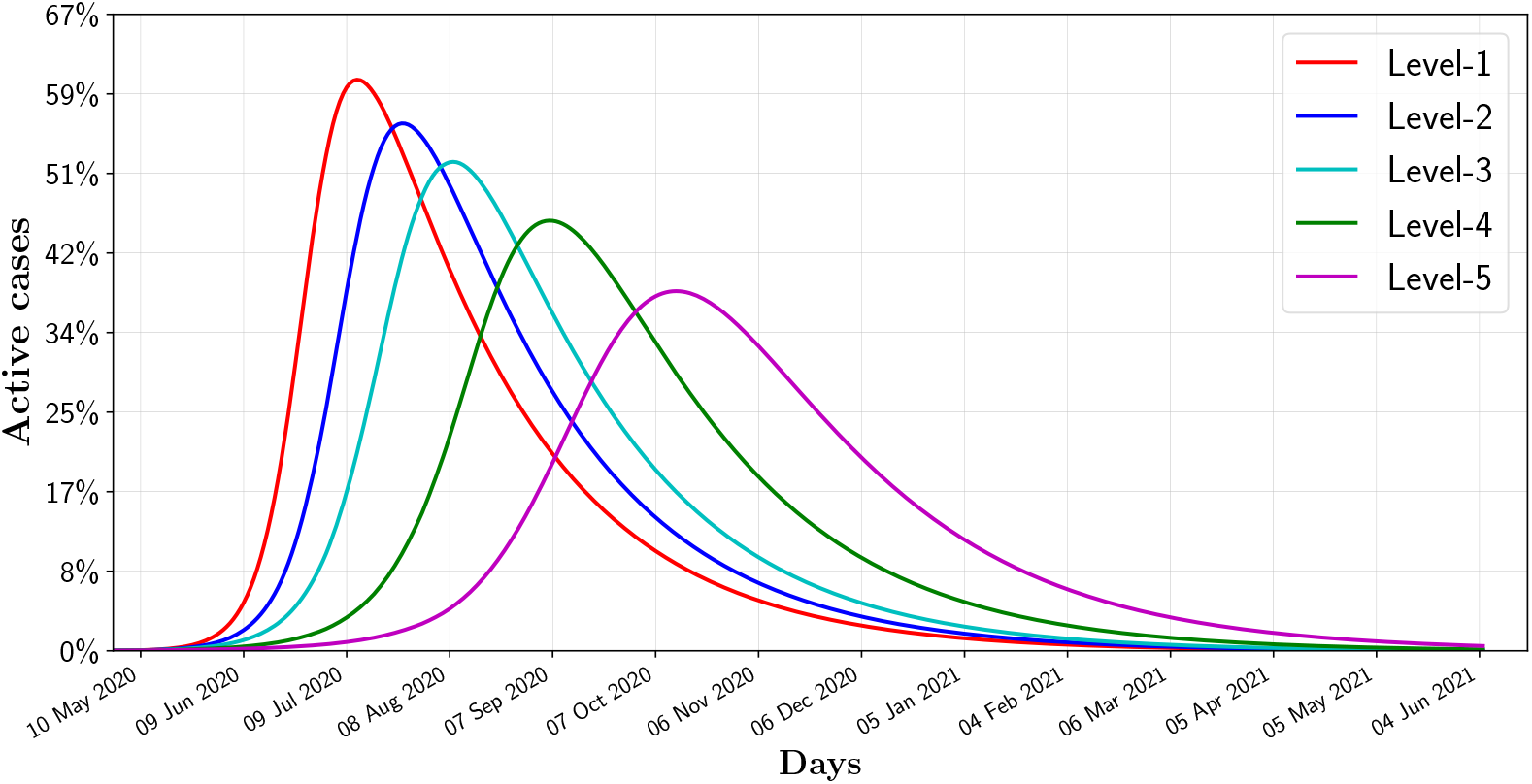
Predictions for South Africa in terms of the differential distribution of active cases for the different alert levels (see text).

One can appreciate the strong effect in terms of the position of the apex that the different restrictions have. This effect becomes more pronounced for smaller deviations from Level 5 restrictions. This speaks to how the outcomes are sensitive to the movement restriction. Level 4 restrictions would effectively delay the peak of the pandemic, where the number of active cases would remain small for about two months followed by rapid growth leading to the peak around September.

Two main sources of uncertainty are considered here. The leading source stems from the variation of the parameter *α*_*s*_. The statistical error of *α*_*s*_ ranges between 2% and 3%. Here a more conservative approach is adopted. The country-to-country variation, which is of order of 10% (68% C.I.), is considered to be a more realistic estimate of the potential deviation from the linear behavior assumed^2^ in Eq. (4). The uncertainty driven by *β*_0_ = 0.28*±*0.003 (68% C.I.) is uncorrelated with the above mentioned uncertainty in *α*_*s*_ and it is also considered here. A sub-leading source of uncertainty is related to the uncertainty in the determination of *γ*_0_ = 0.022 ± 0.001 (68% C.I.), the daily recovery rate. Figures 3 and C.7 illustrate the effect of these uncertainties in regard to predictions. Results in Figures 3 and C.7 are given in terms of the cumulative number of cases, recovered and mortality with the corresponding band. The relative differential distribution for active cases and the symptomatic population are also reported. The format used here is the same as the one used in Figure 2.

**Figure 3:**
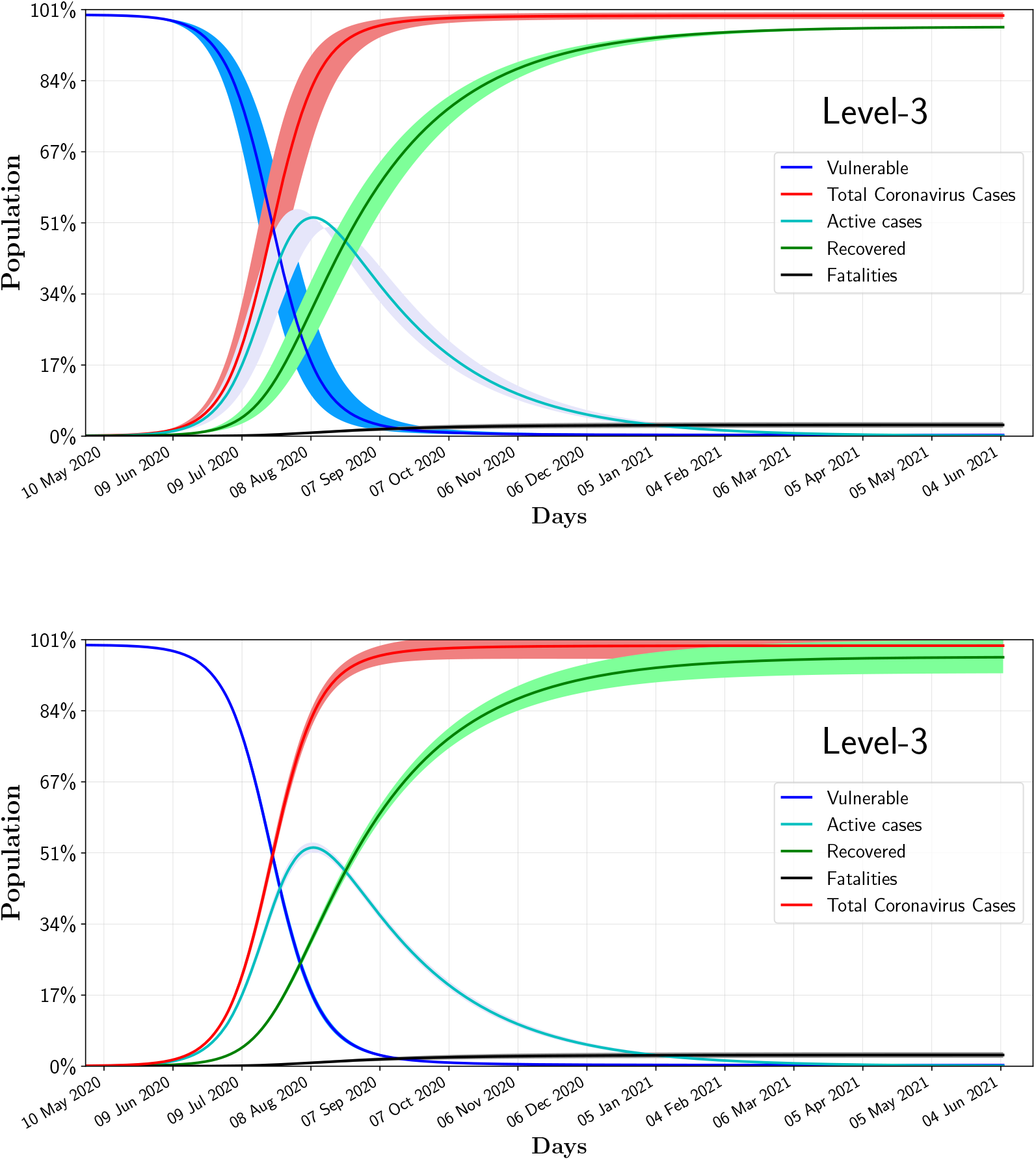
Uncertainties of the model for Level 3 restrictions. The upper and lower graphs correspond to uncertainties in *α*_*s*_, *β*_0_ and *γ*_0_, respectively (see text).

### 4.1. Gauteng Predictions and model re-calibration

The parameters obtained for South Africa, shown in Table 5, are initially applied to the Gauteng Province data to produce estimate predictions for Levels 4 and 5. These predictions are compared to Gauteng’s May data in Figure 4.

**Figure 4:**
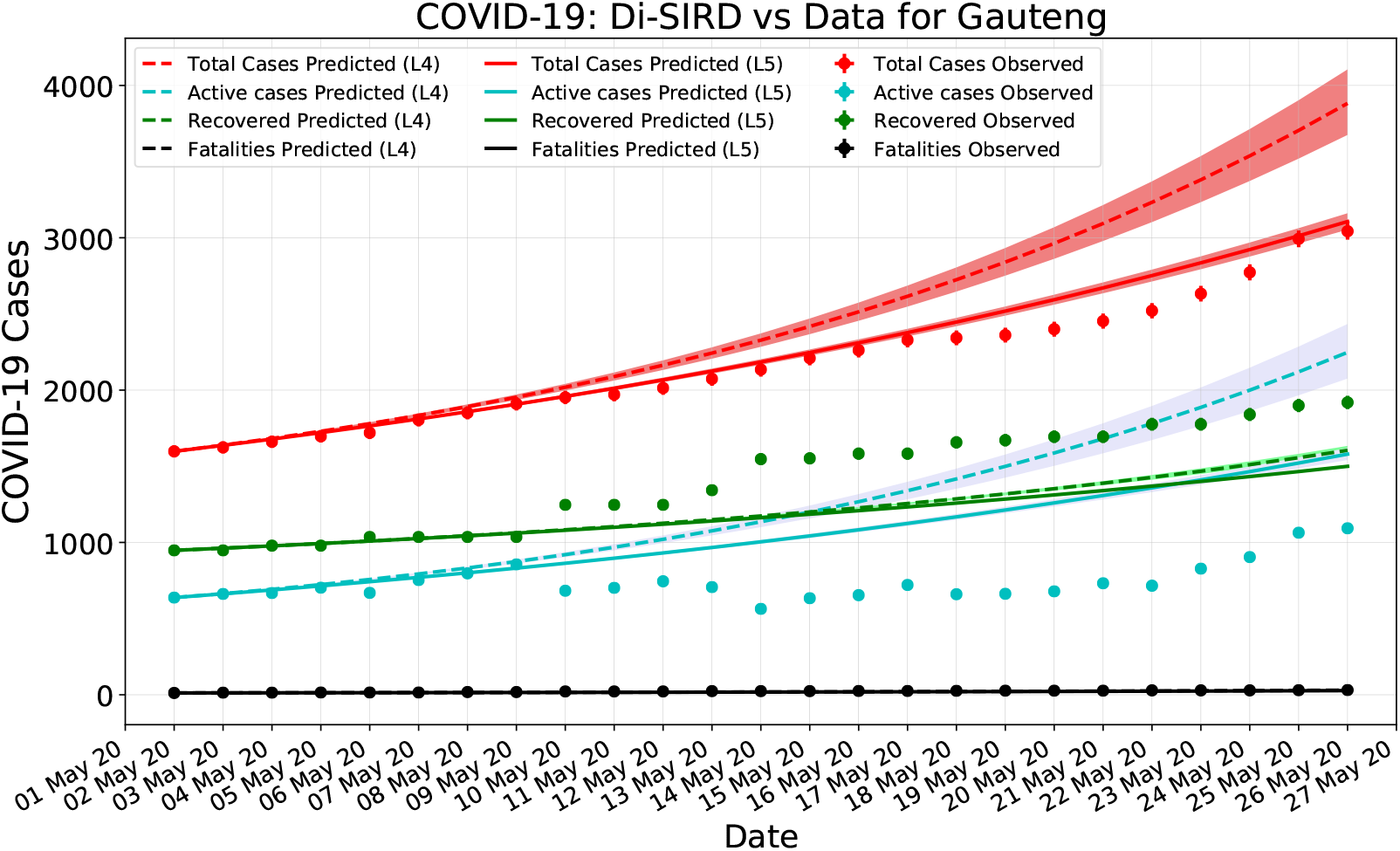
Gauteng province predictions versus data for alert Levels 4 and 5 using the parameters obtained during lockdown (see text).

The discrepancies in the number of active and recovered cases were due to a fluctuation in the number of recoveries that took place in the middle of May. It is relevant to compare the total number of cases as predicted with parameters obtained in Section 4 to the data. One can appreciate that at the total number of cases lie below what would have been the expectation of Level 4 and even Level 5. This is indicative of the fact that adherence to social distancing by the province population increased significantly with respect to what was observed before the lockdown. This is expected, although it is very difficult to predict. Here we adopt a semi-empirical approach where the parameter responsible for the efficiency of the NPIs is re-adjusted with the data in order to make further predictions regarding the evolution of the pandemic.

The *γ* and *α* parameters are re-calibrated for Gauteng Province alert Level 4 data. Towards the end of May the province experienced a significant increase in the number of cases due to the emergence of “hot-spots”, whose dynamics is different and depend strongly on the characteristics of local fluctuations. The dynamics of local fluctuations is not included in the modelling here. As a result, the recovery rate *γ*_*gp*_ is recalculated to include Level 4 data until May 26^*th*^:

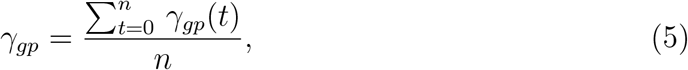

where *t* ranges from March 15^*th*^ to May 26^*th*^ 2020. The *β* equation for recalibration becomes of the form:

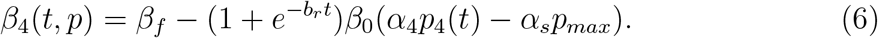

The value of *α*_4_ is then determined through a regression fit on Gauteng’s May data. The re-calibration parameters are shown in Table 6. It is important to note that the efficiency of the NPIs, or the adherence to social distancing, has increased by 20%. Figure 5 shows the alert Level 4 modelling for Gauteng versus data with the re-calibrated parameters applied. Predictions from for the Gauteng province based on this re-calibration procedure are shown in Appendix Appendix C for the different alert levels.

**Table 6:** Parameters obtained with Gauteng’s data up until May 26^*th*^ 2020.

|  |  |
| --- | --- |
| $\gamma_{gp}$ | $\alpha_4$ |
| 0.035 | 0.0089 |

**Figure 5:**
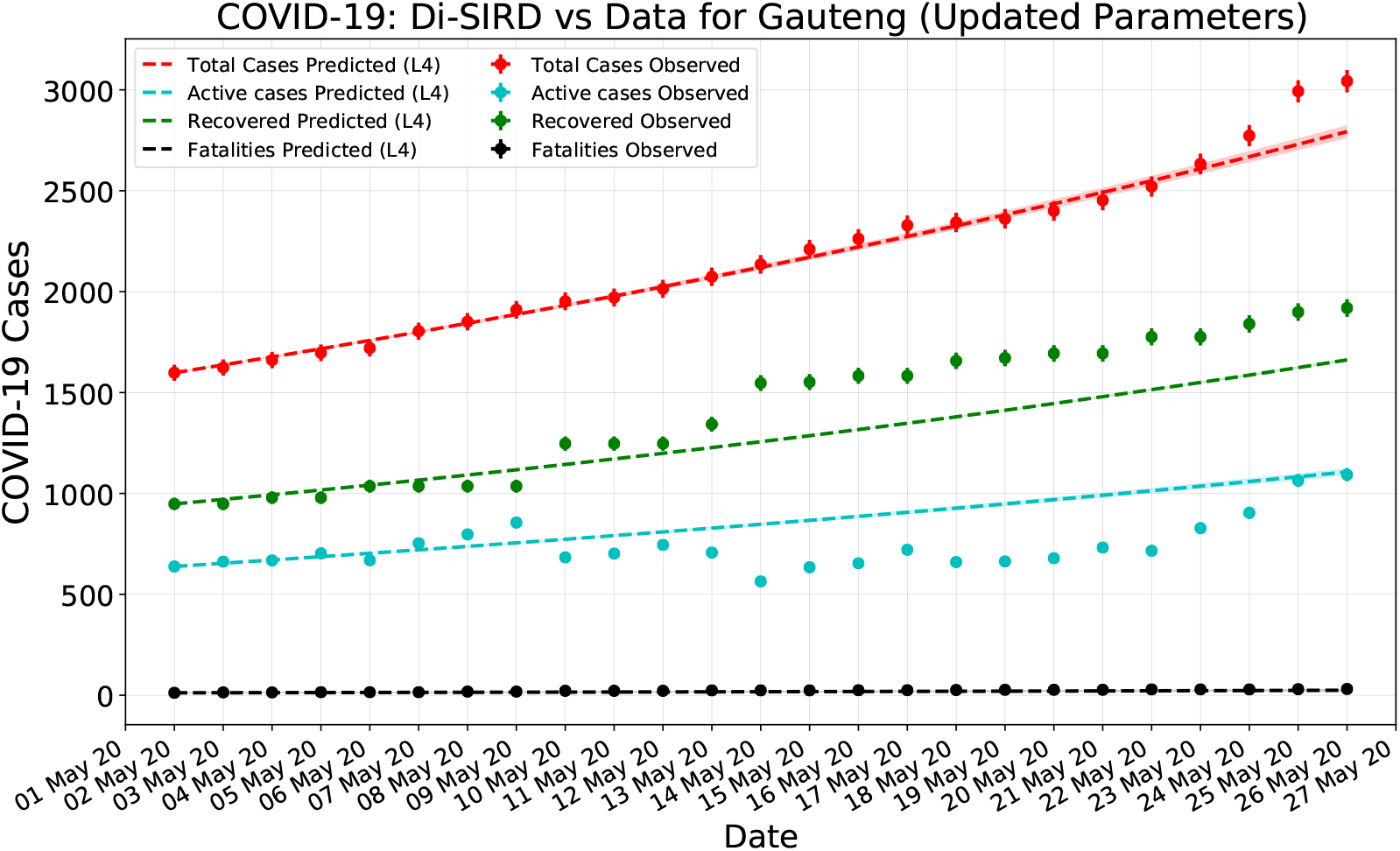
Gauteng province predictions versus data with re-calibrated parameters using Level 4 stringency (see text).

## 5. Discussion and Conclusions

While lockdown measures have been successful in curbing the spread, our study indicates that removing them too swiftly will result in the resurgence of the spread within one to two months. Reducing the stringency index by 10 will delay reaching the apex by about 6 months, where reducing it by 20 will delay by only four months^2^. The amplitude of the apex increases by about a factor of two by moving from Δ*p* = − 10 to Δ*p* = −20.^2^ This indicates that post-lockdown measures should be staged and the reduction of the stringency index should be slow if they are to be applied in a sustained fashion.

All relevant parameters that enter the temporal evolution of the spread need to be tuned such that the expected peak of the number of cases and ICU usage falls below the thresholds characteristic to the country. One can denote a vector of critical parameters for which a country’s healthcare system is not overwhelmed:

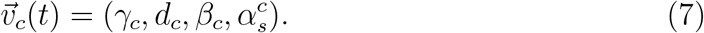

It can be reasoned that *γ*_*c*_ and *d*_*c*_ should display a weak time dependence in that they primarily depend on medical advances, rather than on NPIs. In this setup the condition for the system to remain sub-critical can be expressed as follows:

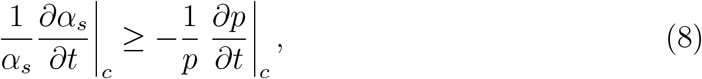

where the temporal partial derivatives are evaluated at the point of criticality defined by 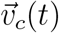.

Given constant *γ* and *d*, it is essential to increase *α*_*s*_. For the system to remain sub-critical, the rate with which *α*_*s*_ increases should outpace that of the decrease of the stringency index. Monitoring of *α*_*s*_ becomes essential to controlling the post-lockdown phase. Equation 8 also serves as a recipe for policy-makers to determine whether an administrative unit is prepared for the transition from one level to another. In particular, Equation 8 supported the transition from alert Level 4 to Level 3 that took place in the province in June 2020.

In the South African case, and given the still relatively low value of *α*_*s*_, it seems evident that it will be difficult to pass from Level 3 to Level 2 in the short to mid term without a significant increase of the efficiency of the NPIs. The data that will be collected in July will be critical to understand the evolution of *α*_*s*_ and its potential enhancement due to adherence to social distancing measures in a system where the stringency index moves relatively slowly.

Given the emergence of “hot-spots” dynamics in South Africa, it is essential to implement local dynamics in the model in order to extract the macroscopic efficiency of NPIs from the data.

## 6. Discussion of Control Strategies

A brief overview of the model developed in the parent paper^2^ is presented here before enumerating on two broad classes of epidemic control strategies which are possible now using our work. The model is depicted in Figure 6 where there are three sub-systems; each of which is coloured differently.

**Figure 6:**
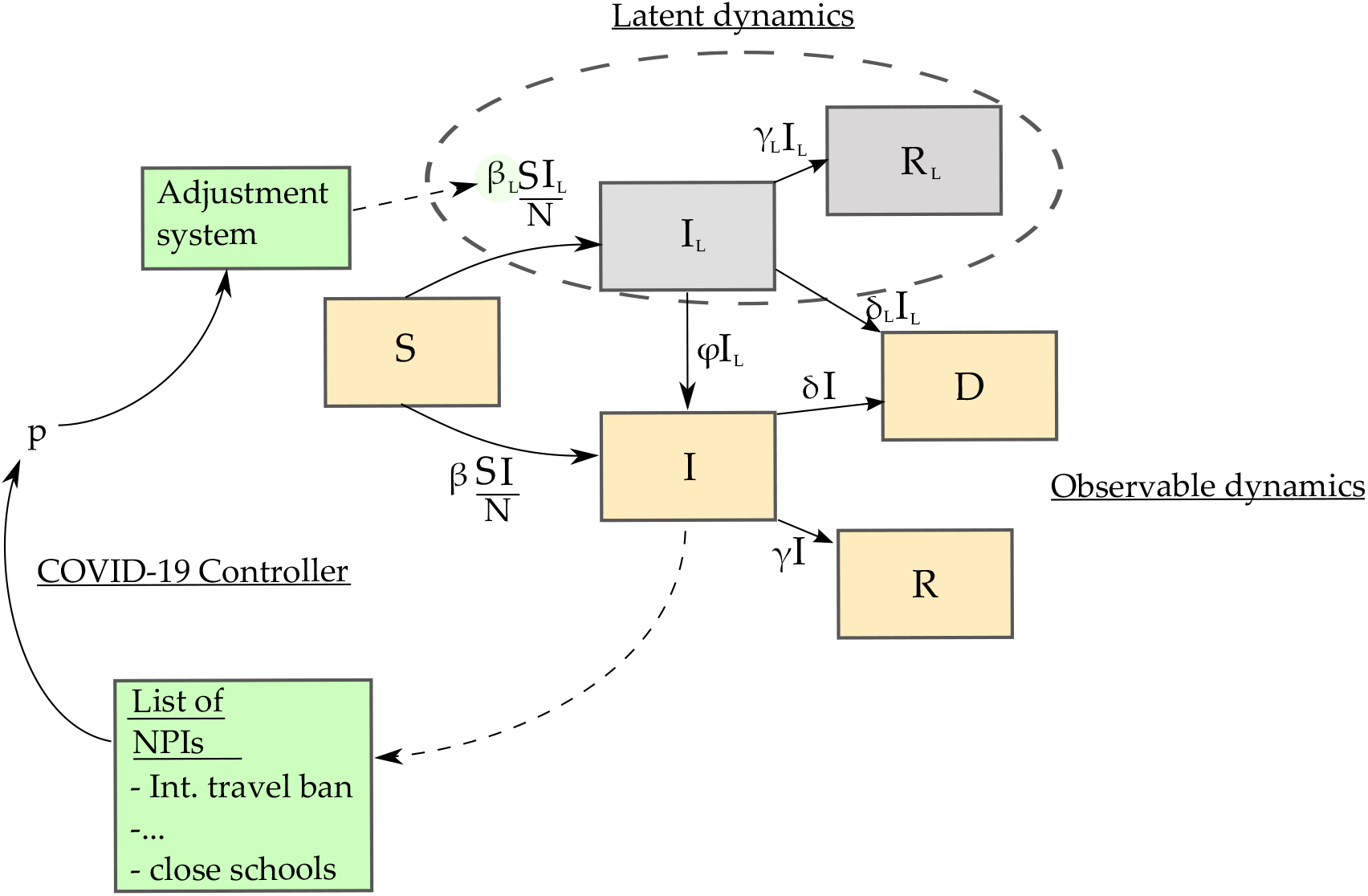
Latent SIRD model with NPI controller of the latent transmission parameter *β*_*L*_. Extended from Choma et al. _2_

The observable dynamics (orange) are what we can actually see during this epidemic; we know the number of people who are susceptible (*S*), the number of observable infected patients (*I*), the number of deaths attributed to COVID-19 (*D*) and the number of recoveries from a known state of infection (*R*). The latent dynamics (grey) are what we cannot see and must infer; these are the latent infections (*I*_*L*_) and the latent recoveries (*R*). The latent dynamics affect the observed dynamics through the *ϕI*_*L*_ term which represents the rate at which hidden infections become known to us. The proposed COVID-19 control system (green) is depicted as receiving information about *I*, activating the level of control from the list of NPIs which in turn implies *p* and this value of *p* filters through the adjustment system to eventually affect *β*_*L*_, the latent infection transmission rate. The adjustment system takes into consideration the typical time it takes to implement and withdraw the NPIs and the level of effect of each NPI for the population under consideration. As such, if the decision maker tries to change the latent infection rate through NPI’s; the results will not be seen immediately and we have quantified this fact within our model.

Control theory has a rich history of successful applications including space flight, computer networking, robotic arm manipulation, automatic military defense systems, magnetic levitation of high speed trains and much else.^8^ This theory has been applied extensively on the present problem of epidemic control. ^9^

There are essentially two options for control given the current NPI framework:

1. Open-loop (time based) control.
2. Closed-loop (infections and time based) control.

There are a myriad of possible control strategies which may be designed for important metrics like, peak infection numbers or time to reach a certain level or total number of deaths etc. A follow up paper is intended to address these rigorously using the models we have developed and offer multiple options with which to augment the current virus containment efforts.

## Data Availability

All data is available on the South Africa Covid-19 Monitoring site: https://www.covid19sa.org/

## 7. Acknowledgements

Authors are indebted to the South African Department of Science and Innovation and the National Research Foundation for different forms of support. This includes, but it is not limited to, support through the SA-CERN Program and the National E-science Postgraduate Teaching and Training Platform. Authors are also grateful for grant support from the IEEE.

## Appendix A

### Rationale for Indicator Scoring

Tables A.7-A.11 outline the rational for the scoring of the S2, S4, S6 and S7 indicators in terms of the South African alert levels.

**Table A.7:** Table Showing Indicator Scoring Rationale for South African **Alert Level 1**.

| OxCGRT Indicator | Value | Rationale |
| --- | --- | --- |
| S2: | 0 | All businesses are allowed to operate. |
| S4: | 0 | All public transport is operational with strict hygiene requirements. |
| S6: | 0. | Inter-provincial travel is allowed. |
| S7: | 0.5 | There is an international travel restriction. |
| **Stringency Level** $36 + (100 * S1/7)$ | | |

**Table A.8:** Table Showing Indicator Scoring Rationale for South African **Alert Level 2**.

| OxCGRT Indicator | Value | Rationale |
| --- | --- | --- |
| S2: | 0.25 | Most businesses are allowed to operate. Some businesses that have a work environment that has a high risk of transmission are not allowed to open. |
| S4: | 0 | All public transport is operational at this level with strict hygiene requirements. |
| S6: | 0.25 | Inter-provincial travel is allowed, with restrictions on travel between level 1 and level 2 areas. |
| S7: | 1 | There is an international travel ban. |
| **Stringency Level** $50 + (100 * S1/7)$ | | |

**Table A.9:**
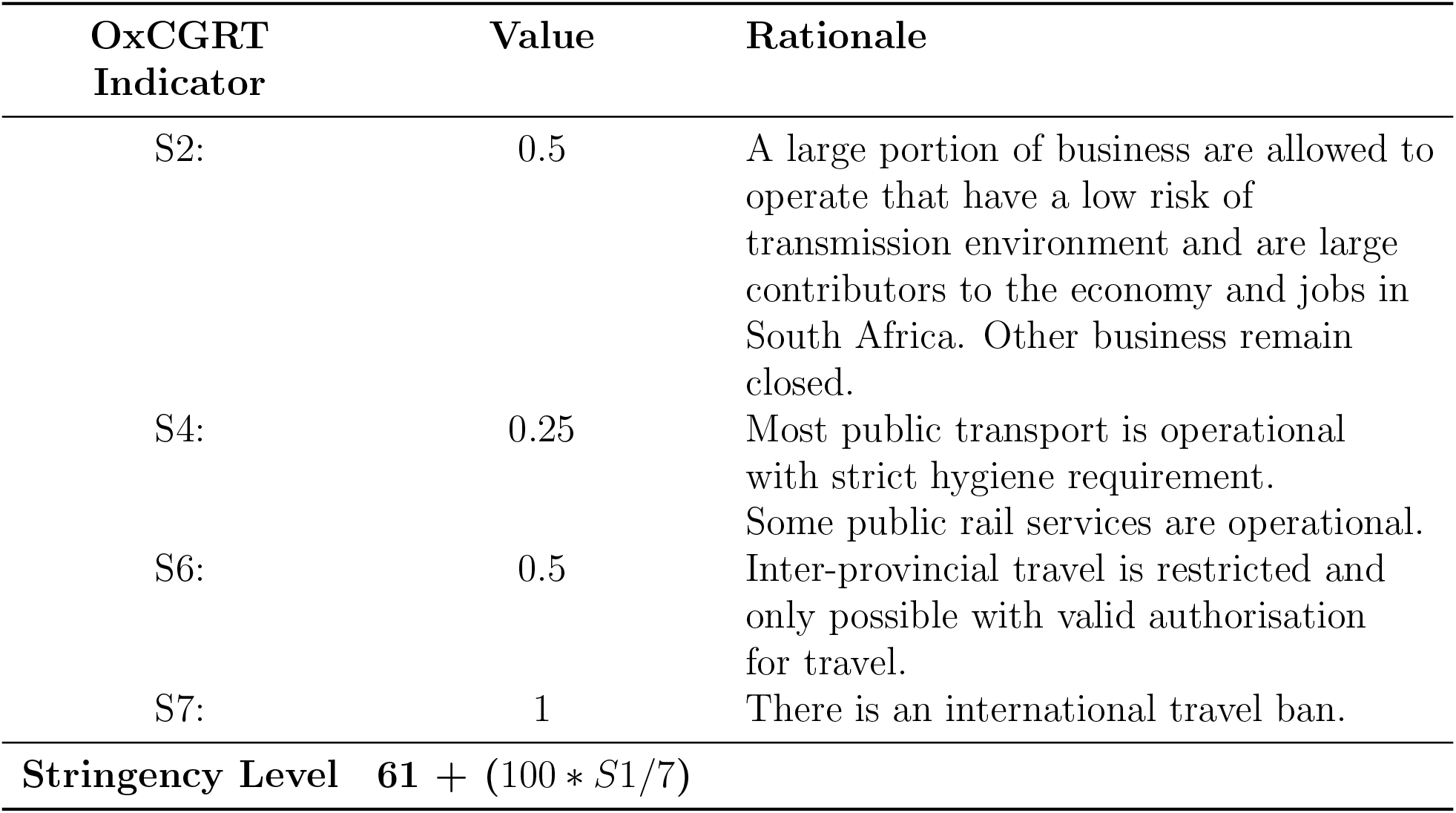
Table Showing Indicator Scoring Rationale for South African **Alert Level 3**.

**Table A.10:** Table Showing Indicator Scoring Rationale for South African **Alert Level 4**.

| OxCGRT Indicator | Value | Rationale |
| --- | --- | --- |
| S2: | 0.75 | A small portion of businesses are allowed to operate that have low risk of transmission environments and are large contributors to the economy and jobs in South Africa. Other business remain closed. |
| S4: | 0.5 | Some public transport is operational at this level with strict hygiene and max capacity regulation. No public rail routes are open. |
| S6: | 1 | Inter-provincial travel is not allowed, except for transportation of goods and exceptional circumstances (e.g. funerals). |
| S7: | 1 | There is an international travel ban. |
| **Stringency Level** $75 + (100 * S1/7)$ | | |

**Table A.11:** Table Showing Indicator Scoring Rationale for South African **Alert Level 5**.

| OxCGRT<br>Indicator | Value | Rationale |
| --- | --- | --- |
| S2: | 1 | All non-essential services are closed. |
| S4: | 1 | All non-essential public movement is banned. Only essential workers may use limited public transport. |
| S6: | 1 | Inter-provincial travel not allowed |
| S7: | 1 | There is an international travel ban. |
| **Stringency Level** $86 + (100 * S1/7)$ | | |

## Appendix B

### Effect of the Varying S1 Indicator

The government announced that the staged reopening of schools will not coincide exactly with the demotion of the alert levels. Instead the staged reopening will commence on the 1 of June 2020. This date is subject to change due to unforeseen circumstances related to the Covid-19 crisis. The staged reopening will be undertaken by periodically allowing strategically paired grades to go back to school in predominantly two week intervals. Numbers were extracted from the “2017 School Realities Report”^10^ for all necessary school level (Pre Gr. R - Gr. 12) student and educator information. The “Statistics on Post-School Education and Training in South Africa 2017”^11^ report was used for all the tertiary numbers for students and staff. Data from 2017 was used instead of 2019 because the 2019 and 2018 statistics report on post-school education have not yet been completed. The *S*1 indicator will represent the portion of total students and staff, in all levels of education in South Africa, that have gone back to work/school. The overall total used for this calculation consists of the following sub-totals:

1. Total number of students in the school system public and private (Pre Gr. R - Gr. 12)
2. Total number of educators in the school system public and private
3. Total number of contact (not distance) students in Higher Education Institutes (HEIs)
4. Total staff of public HEIs
5. Total number of students in private HEIs
6. Total staff in private HEIs
7. Total number of students in Technical and Vocational Education and Training (TVET) colleges
8. Total number of TVET college Staff (estimation based on CET student/staff ratio)
9. Total number of students in Community Education and Training (CET) colleges
10. Total number of CET staff
11. Total number of private college students
12. Total number of private college staff

All required numbers were attainable from the aforementioned sources except for the total number of TVET staff. An estimate for this number was calculated by using the student to staff ratio from the CET Colleges in South Africa.

The chosen grade pairs, the corresponding percentage of total students and staff and the appropriate *S*_1_ equation for different stages of school opening can be seen in Table B.12.

**Table B.12:** Table Showing Indicator Contribution to the Stringency Index for different South African Alert Levels. *Grade Totals Include Educators

| | Population | | Equation for $S_1$ |
| --- | --- | --- | --- |
|  | Grade Totals* | % of Total Persons |  |
| Stage 1: |  |  |  |
| Gr.7 | 924167 | 6.0% | $S_1 = (1 - \frac{10.3}{100})$ |
| & Gr.12 | 661116 | 4.3% |  |
| Total Combined: | 1657888 | 10.3% |  |
| Stage 2: |  |  |  |
| Gr.6 | 978130 | 6.3% | $S_1 = (1 - \frac{10.3+12.1}{100})$ |
| & Gr.11 | 892784 | 5.8% |  |
| Total Combined: | 1935262 | 12.1% |  |
| Stage 3: |  |  |  |
| Gr.5 | 1046370 | 6.8% | $S_1 = (1 - \frac{36.1}{100})$ |
| & Gr.10 | 892784 | 5.8% |  |
| Total Combined: | 2195289 | 13.7% |  |
| Stage 4: |  |  |  |
| Gr.4 | 1130949 | 7.3% | $S_1 = (1 - \frac{49.2}{100})$ |
| & Gr.9 | 894113 | 5.8% |  |
| Total Combined: | 2094711 | 13.1% |  |
| Stage 5: |  |  |  |
| Gr.3 | 1124312 | 7.3% | $S_1 = (1 - \frac{62.8}{100})$ |
| & Gr.8 | 971367 | 6.3% |  |
| Total Combined: | 2167757 | 13.6% |  |
| Stage 6: |  |  |  |
| Gr.2 | 1141325 | 7.4% | $S_1 = (1 - \frac{77.9}{100})$ |
| & Gr.1 | 1186829 | 7.7% |  |
| Total Combined: | 2408228 | 15.1% |  |
| Stage 7: |  |  |  |
| Gr.R | 868389 | 5.4% | $S_1 = (1 - \frac{83.3}{100})$ |
| Total: | 868389 | 5.4% |  |

Table B.13 shows the effect that the S1 indicator has on the Stringency Index for different Alert Levels:

**Table B.13:**
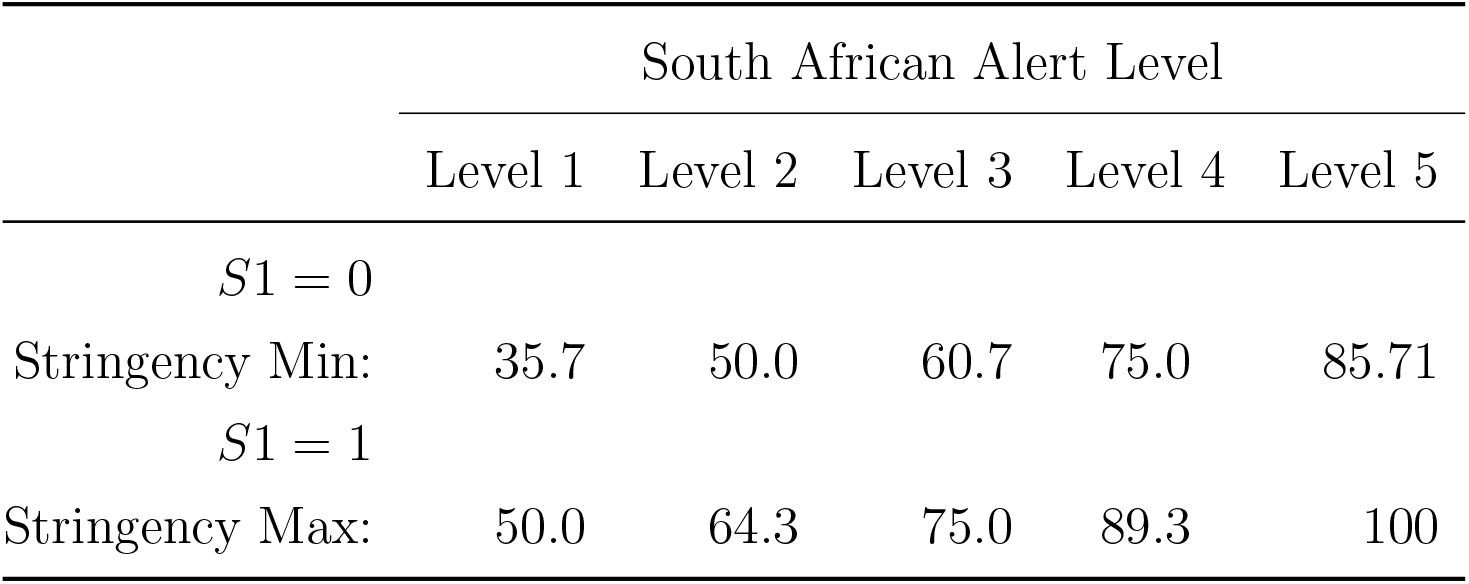
Table Showing Indicator Contribution to the Stringency Index for different South African Alert Levels.

## Appendix C

### Additional Graphs

This appendix contains additional graphs that are referred to in the body of the text. Figure C.7 shows predictions for South Africa for Level 4 restrictions.

**Figure C7:**
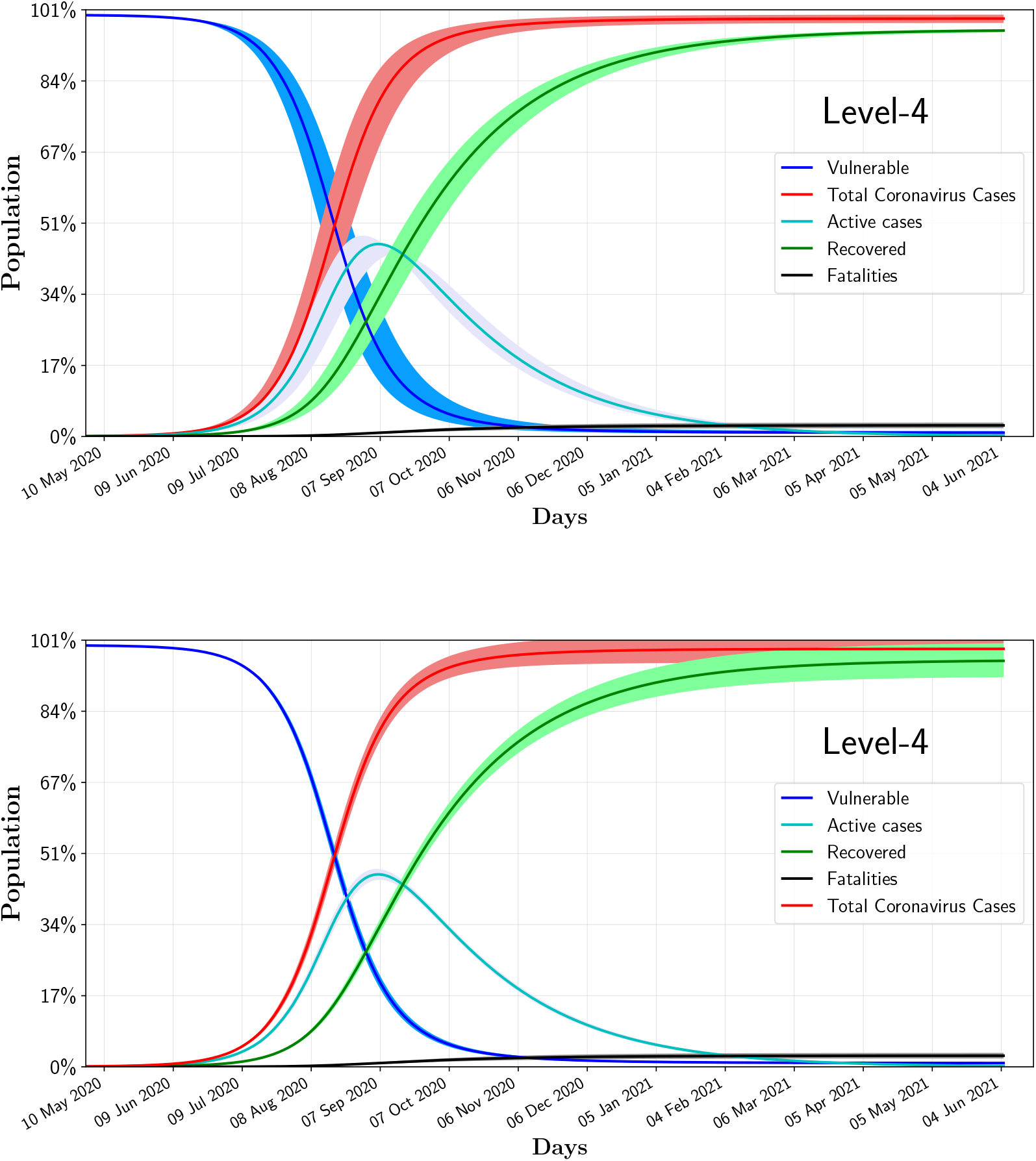
Uncertainties of the model for Level 4 restrictions. The upper and lower graphs correspond to uncertainties in *α*_*s*_ and *γ*_0_, respectively (see text).

Figure C.8 displays the long term predictions for the Gauteng Province with the re-calibration performed with data collected in May, as described in Section 4.1. The graph assumes that 10% of the population will become symptomatic. The graphs assume that no changes in alert level takes place. Currently, the Gauteng province is in alert Level 3.

**Figure C8:**
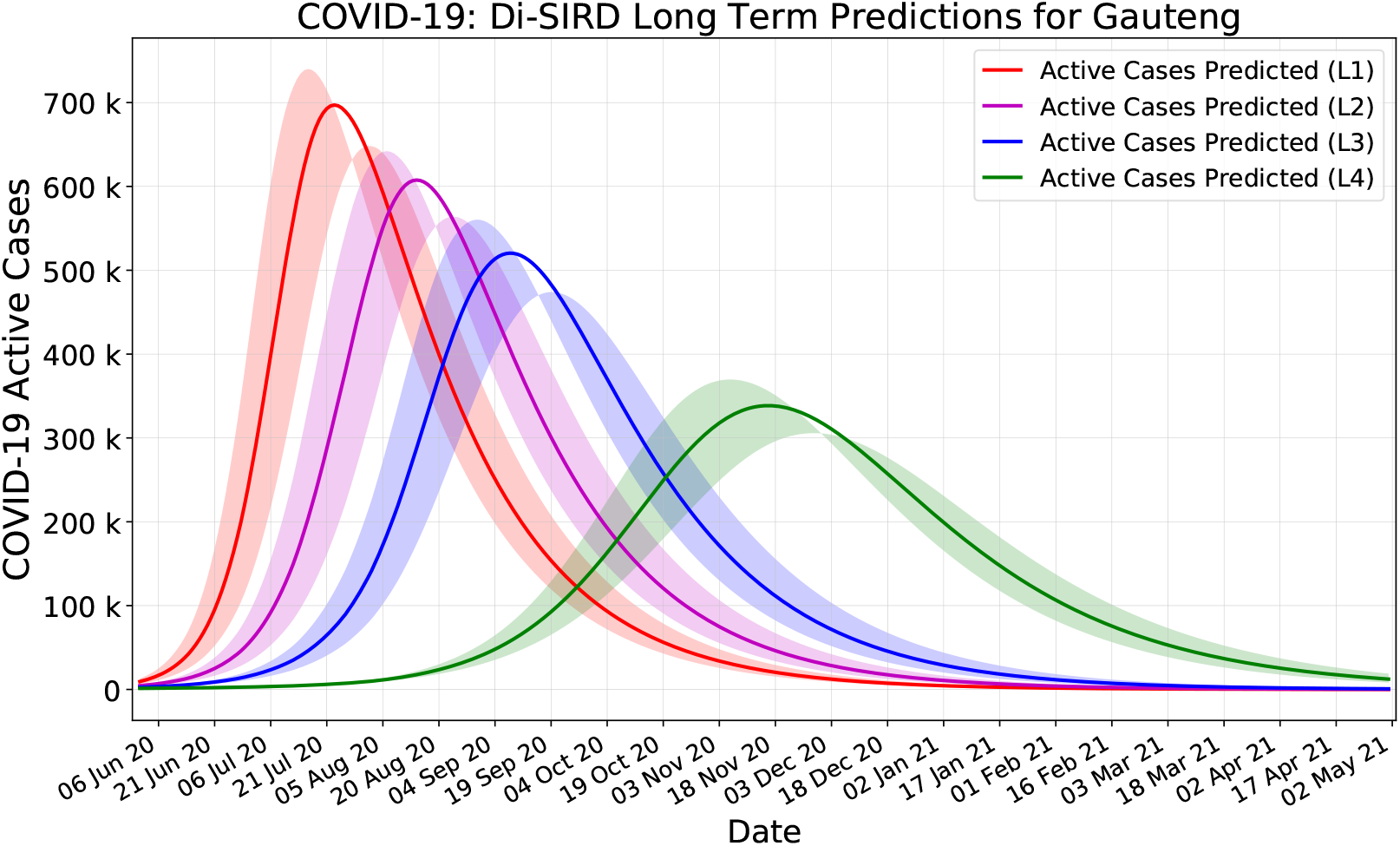
Long term re-calibrated predicted number of active cases in Gauteng Province

